# Analysis of ECMO Outcomes in Patients with Known Adrenal Insufficiency: Insights from the National Inpatient Sample Database

**DOI:** 10.1101/2024.05.21.24307575

**Authors:** Aastha Randhawa, Kundan Jana, Moin Sattar

**Affiliations:** Department of Medicine, Maimonides Medical Center

## Abstract

Adrenal crises occur due to either a complete lack or an insufficient amount of cortisol and are usually triggered by stressors like infections, trauma, surgical interventions, and dental procedures. The physiological changes associated with major surgery are well-studied stressors linked to adrenal crisis. Extracorporeal membrane oxygenation (ECMO), a strategy employed to assist with circulatory support and gas exchange, is also a significant physiologically demanding stressor to the human body. In this study, we retrospectively assess the incidence and outcomes of patients with adrenal insufficiency who were cannulated for extracorporeal membrane oxygenation (ECMO) using the National Inpatient Sample (NIS) database. Patients who were cannulated for ECMO from 2016 to 2020 were identified and divided into two cohorts depending on the presence or absence of adrenal insufficiency. The two groups were compared for the primary outcome of inpatient mortality and secondary outcomes including rates of LVAD, heart transplant, lung transplantation, stroke, GI bleed, and renal failure.

In our study, we found that mortality rates between the two groups were similar. All-cause inpatient mortality was 45% in both groups (p>0.9). We found no differences in the rates of heart transplantation, lung transplantation, or LVAD implantation between patients with and without adrenal insufficiency. We additionally find that mortality rates in our study mirror those from dedicated ECMO registries, further lending credence to our findings. Our study discusses important findings from a relatively understudied disease process, based on a large and representative population from multiple centers across the country.

## Introduction

Adrenal crises occur due to either a complete lack or an insufficient amount of cortisol, the main glucocorticoid of the human body. In these situations, the intrinsic glucocorticoid activity falls short of physiological demands required to maintain homeostatic balance.^1^ This manifests as a vague constellation of symptoms such as hypotension, acute abdominal symptoms or altered mental state. ^2^ Gastroenteritis and fever are the primary triggers in 60–70% of cases, though crises can also be induced by various other stressors, including trauma, surgical interventions, dental procedures, and significant physiological distress. ^1^

Surgical interventions are a well-studied stressor linked to adrenal crisis.^3^ The physiological changes associated with major surgery include inflammatory cascades and fluid shifts as a result of vasodilatation, increased capillary permeability and an impaired response to catecholamines. The general treatment paradigm includes prompt administration of parenteral glucocorticoid along with appropriate fluid resuscitation. Pharmacological supplementation of patients with adrenal insufficiency remains more challenging in those with concomitant long standing cardio-respiratory disease, especially in the setting of acute cardiopulmonary collapse. Such patients can be rescued with invasive mechanical support systems that represent some of the most physiologically demanding stressors to the human body.

Extracorporeal membrane oxygenation (ECMO) is a strategy employed to assist with end-stage respiratory failure, cardiac failure, or both, by utilizing an external circuit for circulatory support and gas exchange. The utilization of ECMO has increased steadily over years. Over the past decade, there has been a remarkable 1,180% increase in the number of adult cardiac ECMO runs, surpassing 2,000, and a substantial 133% rise in the number of ECMO centers. ^4^ Despite this, data on adrenal insufficiency in these settings are limited. Here we retrospectively assess the incidence and outcomes of patients with adrenal insufficiency in patients cannulated for extracorporeal membrane oxygenation (ECMO).

## Methods

This is a retrospective observational study using the National Inpatient Sample (NIS) database. This dataset represents the largest publicly available all-payer inpatient healthcare database designed to produce U.S. regional and national estimates of inpatient utilization, access, cost, quality, and outcomes.^5^

We identified patients who were cannulated for ECMO from 2016 to 2020 using the following ICD10 codes: ESA002, 5A15223, 5A1522F, 5A1522G, 5A1522H, 5A15A2F, 5A15A2G, 5A15A2H. Out of these, patients were divided into two cohorts depending on the presence or absence of adrenal insufficiency. The diagnosis of adrenal insufficiency included all ICD-10 codes for primary, secondary or tertiary adrenal insufficiency, irrespective of etiology. The following ICD10 codes were utilized: E271, E272, E273, E2740, E2749, E279, E250, E71511, E71518, E71520, E71521, E71522, E71528, E71529, A187.

Comorbidities such as Hypertension, Heart failure, Chronic respiratory failure, Diabetes, Obesity were also accounted for. Finally, the two groups were compared for the primary outcome of inpatient mortality and secondary outcomes including rates of LVAD, Heart Transplant, Lung Transplantation, Stroke, GI Bleed, and Renal failure. This study is IRB exempt, given the NIS uses anonymized public-use data without personally identifiable information.

Comparison of continuous variables were performed via the Wilcox Rank-Sum test, and categorical variables were studied via the Pearson’s Chi-squared tests wherever appropriate. Remaining sub-analyses were performed without a-priori power analyses. Statistical analyses were performed using R (v4.1.0).

## Results

We identified 10,910 patients who were cannulated for ECMO between 2016 and 2020 from the NIS database. 292 of these patients had concomitant adrenal insufficiency (AI), the remaining 10618 patients were used as controls.

Baseline characteristics are shown in Table 1. Patients with AI were younger (median 48 years vs 55 years; p < 0.001), and more likely to be women (43% vs 36%; p = 0.014). Notably patients with AI were also less likely to have comorbidities such as Hypertension or Diabetes (p < 0.001).

**Table 1:** Baseline characters of patients in the AI and Control groups.

| Cohorts: | Control, N = 10,618<sup>1</sup> | Adrenal Insufficiency, N = 292<sup>1</sup> | p-value<sup>2</sup> |
| --- | --- | --- | --- |
| Patient Age (years) | 55 (40, 64) | 48 (32, 60) | <0.001 |
| Sex |  |  | 0.014 |
| Female | 3,835 (36%) | 126 (43%) |  |
| Male | 6,782 (64%) | 166 (57%) |  |
| Unknown | < 10 | < 10 |  |
| Hypertension | 5,496 (52%) | 122 (42%) | <0.001 |
| Diabetes | 2,634 (25%) | 45 (15%) | <0.001 |

<sup>1</sup> Median (IQR); n (%)
<sup>2</sup> Wilcoxon rank sum test; Pearson's Chi-squared test

On univariate analyses, we show that there were no significant differences in inpatient mortality (primary outcome) between the two groups (Table 2), and no significant differences in the rates of inpatient LVAD, Heart Transplant, or Lung Transplantation. We studied secondary outcomes of stroke, GI Bleeds, and Renal failure, and found no statistically significant differences between patients on ECMO with and without AI.

**Table 2:** Outcomes of patients in the AI and Control groups.

| Cohorts: | Control, N = 10,618<sup>1</sup> | Adrenal Insufficiency, N = 292<sup>1</sup> | p-value<sup>2</sup> |
| --- | --- | --- | --- |
| Inpatient Mortality | 4,734 (45%) | 130 (45%) | >0.9 |
| LVAD | 560 (5.3%) | 10 (3.4%) | 0.2 |
| Heart Transplant | 274 (2.6%) | < 10 (3.1%) | 0.6 |
| Lung Transplant | 663 (6.2%) | 17 (5.8%) | 0.8 |
| Stroke | 1,280 (12%) | 42 (14%) | 0.2 |
| GI Bleed | 1,019 (9.6%) | 37 (13%) | 0.08 |
| Renal Failure | 7,276 (69%) | 210 (72%) | 0.2 |

<sup>1</sup> n (%)
<sup>2</sup> Pearson's Chi-squared test

On multi-variate analyses performed to assess the confounding effects of baseline differences noted above, we found no significant differences in mortality based on the incidence of adrenal insufficiency except for the effect of patient age. Age at the time of cannulation for ECMO was significantly and independently associated to mortality (p < 0.001).

We performed additional sub-analyses and found no statistically significant differences in post-heart transplant and post-LVAD mortality within the index hospital admission. Post-lung transplant mortality with adrenal insufficiency was however significantly worse (9.9% vs 1.8% in controls), however these results are exploratory and likely too underpowered to draw reliable conclusions from.

## Discussion

Adrenal insufficiency has been vaguely associated with excess morbidity, mortality, and impaired quality of life. ^6–8^ Most prior studies were performed in the outpatient setting and are less relevant for inpatient management.

Patients with AI are at risk of life-threatening adrenal crisis due to their inability to compensate in periods of stress by increasing glucocorticoid production. The body normally responds to significant stressors, such as major surgery, by increasing circulating cortisol levels by 5 to 10 times baseline levels. Data shows that identifying patients with possible adrenal insufficiency and administering appropriate glucocorticoids in periods of anticipated surgical stress can improve outcomes. The recommendations regarding timing and dosage of glucocorticoids are debated.^9^ Proposed recommendations were derived on the invasiveness of surgery and a baseline suspicion of hypothalamic-pituitary axis suppression rather than true randomized control trials.^10–12^

In contemporary literature, the standardized mortality rates for patients with both primary and secondary adrenal insufficiency are more than twice those for a control population adjusted for age and sex. ^6^ Patients with secondary adrenal insufficiency (SAI) also exhibited elevated cardiovascular mortality. ^13–15^ In our study, we see that mortality rates between the two groups were similar. All cause in-patient mortality is seen to be 45% in both groups (p>0.9). These figures are similar to literature on patients undergoing Veno-Venous (VV) and venoarterial (VA) ECMO, with inpatient mortality rates between 30-40% and 60-70% respectively. ^16,17^

Prior studies have shown similar mortality rates amongst male and female patients.^18^ Our study shows that at least on multi-variate analyses gender was not an independent predictor of mortality. We do however note that those with adrenal insufficiency in this cohort were more likely to be women. This observation of a female predominance in adrenal insufficiency aligns with the work of others. ^19^

One of the key factors influencing these outcomes are pre-existing comorbidities. ^20^ Prior studies have shown that diabetes is a risk factor for mortality in patients who undergo VA-ECMO.^21^ A multicentric prospective study conducted during the COVID pandemic revealed significantly higher mortality rates in patients who were either older (age > 60), or had pre-existing comorbidities such as hypertension and renal failure. ^22^ We studied the contribution of hypertension and diabetes given the differences noted at baseline between the adrenal insufficiency and control groups. On multivariate analyses, these were not significantly associated with mortality.

Common non-procedural complications of ECMO are hemorrhage (gastrointestinal or retroperitoneal), thrombosis, stroke, and infection.^22,23^The incidence of bleeding is estimated at 30% to 60% of patients, varying based on the ECMO cannulation strategy and the reason for its use. ^4^ Gastrointestinal bleeding, specifically, occurs in 4.5% to 15.6% of patients on ECMO.^24^ In our study the rates of gastrointestinal bleed was noted to be 9.6% in control group and 13% in study group. Although the difference was not statistically significant, the use of corticosteroids is a known risk factor for GI bleed. It is plausible that patients with known adrenal insufficiency may have received higher doses of corticosteroids while they were being supported on ECMO.^25^ Neurological injury frequently leads to mortality and morbidity.^26^ In studies in children, 7.4% of patients treated with ECMO experienced i’1ntracranial hemorrhage, while 5.7% of all ECMO-treated patients suffered from Cerebral infarction. ^27^ Our results reveal a 12% incidence of stroke in the control group and 14% in the group with adrenal insufficiency. While these numbers are higher than those observed in previous studies, it is possible that coding errors may have failed to distinguish between strokes occurring before or after initiation of ECMO.

The success of ECMO ultimately depends on a successful transition away from temporary mechanical support. This can either be a bridge to recovery, a lung or heart transplant, a left ventricular assist device (LVAD), or a total artificial heart.^20^ In our study we found no differences in the rates of heart transplantation, lung transplantation, or (LVAD) implantation between patients with adrenal insufficiency and without. While our study is not powered to study post-transplantation outcomes, we note that inpatient mortality rates of patients who received either heart transplants or LVADs were similar regardless of whether they had adrenal insufficiency. Outcomes post lung-transplantation however were significantly worse in patients with adrenal insufficiency on univariate analyses.

There were some limitations in our study. While the NIS dataset is a good refection on healthcare trends and outcomes, it includes only ICD-10 billing diagnosis, and lacks data such as laboratory values, patient history, and pharmacy information. For example, in our study, it was impossible to establish whether patients received appropriate doses of corticosteroid therapy, which could have influenced outcomes. Furthermore, the chronology of events can only be assumed. It cannot be established with certainty whether a particular diagnosis was given before or after ECMO was initiated. In our study it is assumed that the diagnosis of adrenal insufficiency must have been provided before or at the outset of an admission. Inadvertent errors relating to billing and documentation may have under or overestimated the prevalence of AI. Finally, the NIS dataset does not offer specific criteria or the clinical indications for initiation of ECMO apart from the fact that it was initiated.

Despite these limitations, our study discusses important findings from a relatively under-studied disease process, from a large and representational population from multiple centers across the country. We additionally find that mortality rates in our study mirror those from dedicated ECMO registries, further lending credence to our findings. Scoring systems for ECMO candidacy selection have been described that take into account comorbidities such as Immunocompromised status, chronic respiratory failure, chronic liver failure, prior cerebrovascular insufficiency and chronic renal failure.^23,28^ There is limited data available on ECMO in patients with pre-existing adrenal insufficiency. As of now, predominant vasoplegic shock resulting from sepsis remains an absolute contraindication for ECMO. Adrenal crisis is known to mimic undifferentiated shock and can often be misdiagnosed in critical care settings. Additional research is required to enhance our understanding of the indications, risks, and potential management considerations for patients with adrenal insufficiency who require ECMO.

## Data Availability

NIS data are available online at the official website of HCUP.

https://hcup-us.ahrq.gov

